# Duodenal alpha-Synuclein pathology and enteric gliosis in advanced Parkinson’s Disease patients

**DOI:** 10.1101/2022.06.16.22276347

**Authors:** Aron Emmi, Michele Sandre, Francesco Paolo Russo, Giulia Tombesi, Federica Garrì, Marta Campagnolo, Miryam Carecchio, Roberta Biundo, Gaya Spolverato, Veronica Macchi, Edoardo Savarino, Fabio Farinati, Piero Parchi, Andrea Porzionato, Luigi Bubacco, Raffaele De Caro, Gabor G Kovacs, Angelo Antonini

## Abstract

**Background:** The role of the gut-brain axis has been recently highlighted as a major contributor to Parkinson’s Disease (PD) physiopathology, with numerous studies investigating bidirectional transmission of pathological protein aggregates, such as α-Synuclein (αSyn), in the context of neurodegenerative disease. However, little and often conflictual evidence is available from human studies due to patient heterogeneity, sampling variability, and the employment of antibodies with affinity for different αSyn epitopes.

**Objectives:** We aimed to investigate the enteric nervous system (ENS) in PD by characterizing αSyn alterations and glial responses in duodenum biopsies of PD patients by employing topography-specific sampling and conformation-specific αSyn antibodies.

**Methods:** 18 Patients with symptomatic PD who underwent Duodopa Percutaneous Endoscopic Gastrostomy and Jejunal Tube (PEG-J) procedure, as well as 18 age- and -sex-matched Healthy Control subjects undergoing routine diagnostic endoscopy, were included in the study. A mean of 4 duodenal wall biopsies were sampled from each patient. Immunohistochemistry was performed for anti-aggregated αSyn (5G4) and GFAP antibodies. Morphometrical-semi-quantitative analysis was performed to characterize αSyn-5G4+ and GFAP+ density and size.

**Results:** Elevated immunoreactivity for aggregated α-Syn was identified in all biopsies of PD patients compared to controls. αSyn-5G4+ colocalized with neuronal marker β-III-tubulin. Evaluation of enteric glia cells revealed an increased size and density when compared with controls, suggesting reactive gliosis.

**Conclusions:** Duodenal biopsy may represent a feasible and reliable tool for characterizing PD pathology in the GI tract and discerning patients from controls. Future studies are required to confirm these findings in a prodromal or early PD phase.

## INTRODUCTION

In Parkinson’s Disease (PD), the role of the gut-brain axis has been greatly highlighted by recent developments in both clinical and preclinical research^1-5^.

There is growing interest in the detection of α-Synuclein (αSyn) deposition, the histological hallmark of PD, in peripheral tissues^6^ including the gastrointestinal (GI) tract^7-10^. Considering the involvement of the enteric nervous system (ENS) in the prodromal stages of PD and its relationship with gut motility^11 12^, the detection of αSyn aggregation and its deposition in gut tissues is of great relevance^13^. Recent animal models suggested the possibility of a bidirectional transmission of αSyn pathology, that may originate either in the enteric nervous system and spread to the brain, or in the amygdala and then move caudally; this has led to the debate on whether the direction of αSyn spreading may impact on disease phenotype and has driven the discussion to the so-called “gut-brain axis”^2 14 15^. These findings have been supported by human postmortem samples^10^, while in-vivo human studies reported heterogeneous immunohistochemistry staining in GI biopsies (mainly gastric and colonic) ^16-18^. Phosphorylated αSyn (p-αSyn) at Serine 129 residue, has been considered as the most reliable marker to distinguish pathological deposits from physiological protein. However, a consensus has not been reached so far on which antibody (anti-αSyn or anti-p-αSyn) should be used in peripheral tissues to distinguish PD patients from controls^9 19 20^, while antibodies specific for αSyn aggregates have been employed in a single study on colonic mucosa, with promising results^21^. Specifically, clone 5G4, the antibody used in this work can detect αSyn aggregates and it was raised against the sequence encompassing aminoacids 46-53 of αSyn^22 23^. Recently, in an in-vitro comparative analysis of several αSyn targeting antibodies, αSyn-5G4 showed high conformational specificity and strong immunoreactivity for all forms of αSyn aggregates with no reaction toward αSyn monomers^24^. Furthermore, αSyn-5G4 immunohistochemistry was more reliable in identifying αSyn aggregates across synucleinopathies compared to other αSyn antibodies and was also able to detect astrocytic and oligodenroglial αSyn inclusions in Lewy Body disease^23 25^.

Furthermore, considerable attention has been drawn to enteric glial cells (EGCs), which may play a critical role in the crosstalk between inflammation and neurodegeneration. According to available studies, EGCs participate in the regulation of gastrointestinal functions, playing a key role in the pathophysiology of gastrointestinal disorders^26 27^. More recently, EGCs have emerged as critical players in regulating GI function in PD, as higher levels of expression for both GFAP and Sox-10, but not of S100-beta, were reported in the GI tract of PD patients. However, levels of glial markers were negatively related to PD disease duration, suggesting that EGC reaction is more relevant at disease onset and decreases over time^28-30^, but the precise mechanism by which EGCs contribute to PD pathogenesis remains to be elucidated.

In the present study we aimed to investigate the histopathological changes in the enteric nervous system by characterizing both αSyn aggregates and enteric glial responses in duodenum biopsies of advanced PD patients with extensive clinical and demographical documentation.

## METHODS

### Subjects

Eighteen (18) patients (12 male, 6 female; mean age 65.2 years, 95% CI 61.4 to 69.0; mean disease duration 11.3 years, 95% CI 9.0 to 13.6) with advanced PD who required initiation of Levodopa Carbidopa Intestinal Gel (LCIG) infusion were part of the study^31^.

All patients underwent Percutaneous Endoscopic Gastrostomy with Jejunal extension (PEG-J) placement; an average of four 3 mm^3^ duodenal-wall biopsies were sampled in a topographically unrelated district to PEG-J placement. Along with the routine clinical assessment (MDS-UPDRS I-II-III, IV, Hoehn and Yahr scales), the Wexner Constipation Score (WCS)^32^ was also calculated (Supplementary Table 1).

Duodenal biopsies from 18 subjects comparable for age- and sex- (9 male, 9 females; mean age 68.6 years, 95% CI 63.8 to 73.4) undergoing screening diagnostic endoscopy were included as healthy controls (HCs). Of note, control subjects were further evaluated clinically and interviewed to exclude any manifestation suggestive of PD or any other neurological disorder.

The study protocol received approval by the ethical committee for clinical experimentation of Padua Province (Prot. n. 0034435, 08/06/2020). Informed consent for the use of biological samples was obtained from all patients. All procedures on human tissue samples were carried out in accordance with the Declaration of Helsinki.

### Tissue processing and staining

Tissue samples were fixed in phosphate-buffered 4% paraformaldehyde, embedded in paraffin, and sectioned at the microtome (5µm slices). Immunoperoxidase staining for aggregated αSyn (Monoclonal Mouse, Clone 5G4, Millipore) and Glial Fibrillary Acidic Protein (GFAP, Monoclonal Rabbit, Dako Omnis) was performed on a Dako EnVision Autostainer station according to manufacturer’s recommendations. Antigen retrieval was performed on a PT-Link Dako Antigen retrieval station using Citrate-buffer at pH 6 solution at 96° for 15 minutes, followed by 1 minute 95% formic acid for the αSyn-5G4 antibody.

Immunoperoxidase staining was repeated at least three times to assure reaction consistency and was independently evaluated by three morphologists blind to the clinical findings. Controversies were resolved by consensus.

### Morphometrical quantification

Photomicrographs were acquired under a Leica DM4500B microscope (Leica Microsystems) connected to a Leica DFC320 high-resolution digital camera (Leica Microsystems) and a computer equipped with software for image acquisition (QWin, Leica Microsystems) and analysis (ImageJ)^33 34^. Whole section photomicrographs were acquired at 5x magnification, while an average of four 20x magnification photomicrographs per available sample were acquired as counting fields and loaded into ImageJ software for semi-automatic immunoreactivity quantification.

The area of the sections was quantified by manually drawing the boundaries of the specimens. A Maximum Entropy Threshold was applied and manually adjusted for each section to discern immunopositive elements from background and negative tissue. Quality control of the applied threshold was performed by an expert morphologist by overlying the thresholded images to the original photomicrographs. Particle analysis was employed with a 0-infinity px threshold to define immunopositive elements quantity and total area occupied within the digital image. For GFAP staining, the number of immunopositive elements was divided by the total area of the sample to obtain a semi-quantitative measure of immunoreactive density within each section. For αSyn-5G4 staining, as immunoreactive structures did not present as distinct elements with defined boundaries, total immunoreactive area (µm^2^) and % of immunoreactive area (A%) was estimated per counting field. Counting fields for each available sample were treated as repeated measures and averaged per single subject.

### Immunofluorescence and confocal microscopy

Fluorescent immunohistochemistry was performed manually. Antigen retrieval was performed on de-paraffinized tissue as in immunoperoxidase staining methods. Following autofluorescence was quenched with a 50 mM NH_4_Cl solution for 10 minutes. Sections were treated with permeabilization and blocking solution (15% vol/vol Goat Serum, 2% wt/vol BSA, 0.25% wt/vol gelatin, 0.2% wt/vol glycine in PBS) containing 0.5% Triton X100 for 90 minutes before primary antibodies incubation. Primary antibodies were diluted in blocking solution and incubated at 4°C overnight. Alexa-Fluor plus 488 Goat antiMouse secondary antibody (Code number: 183 A32723) and Alexa-Fluor plus 568 antiRabbit secondary antibody (Code number: A-11011) were diluted 1:200 in blocking solution as above and incubated for 60 minutes at room temperature. Hoechst 33258 were used for nuclear staining (Invitrogen, dilution: 1:10000 in PBS) for 10 minutes. Slides were mounted and coverslipped with Mowiol solution. Confocal immunofluorescence z-stack images were acquired on a Leica SP5 Laser Scanning Confocal Microscope using a HC PL FLUOTAR 20x/0.50 Dry or HCX PL APO lambda blue 40X/1.40 Oil objectives. Images were acquired at a 16-bit intensity resolution over 2048 × 2048 pixels. Z-stacks images were converted into digital maximum intensity z-projections, processed, and analyzed using ImageJ software.

The antibodies used for IF were the following: mouse anti-aggregated αSyn clone 5G4 (MABN389, Sigma-Aldrich, 1:1000); rabbit Glial Fibrillary Acidic Protein (GFAP, Dako Omnis, 1:1000); mouse β-III Tubulin (#T8578 Sigma; 1:300).

### Statistical Analyses

Statistical analyses and visualizations were performed using GraphPad Prism v.9. Nonparametric data were analyzed with Mann–Whitney U-test. Pearson’s correlation analysis has been employed to assess possible correlations between αSyn expression in duodenum and clinical characteristics, including motor and non-motor scales, cognitive assessments and main non-motor symptoms of PD. Values are indicated as the median, with significance as follows: *\*p <0.05, **p < 0.01, ***p < 0.001*, and *\*\*\*\*p < 0.0001*.

## RESULTS

### Alpha-Synuclein Pathology

αSyn-5G4 immunoreactive elements detected in duodenal specimens were classified according to morphological criteria into four groups: i) Compact and globular immunoreactivities (Figure 1A, B); ii) granular cellular immunoreactivities (Figure 1C, D), mostly resembling cross-reactivity with resident mast cells; and iii) dot-like immunoreactivities (Figure 1E, F), which we interpreted as either crossreaction with lipopigment staining in the duodenal mucosa, or as actual aggregated αSyn deposits if colocalizing with neuronal structures via immunofluorescent staining / double label immunoperoxidase staining. These three morphologies were found in both PD and HC. The fourth morphology was observed as threaded immunoreactivities (Figure 1G, H), which represented the most reliable immunoreactivity type to discern PD from controls. Immunofluorescent staining (Figure 1I-I2) confirmed the colocalization between αSyn-5G4 threaded immunoreactivities and β-III Tubulin, a pan-neuronal and neuritic marker, indicating aggregated αSyn deposits in duodenal nerve fibers of the mucosa and submucosa. Regardless of morphology, immunoreactivity for aggregated αSyn was absent (4/18) or barely detectable (14/18) in controls (Figure 1 L-M), while all duodenal samples collected from PD patients were characterized by marked (18/18; 100%) immunoreactivity for aggregated αSyn (Figure 1N-O). Semi-automatic morphometric quantification for the burden of aggregated αSyn immunoreactivity revealed statistically significant higher immunoreactive tissue area in PD patients compared to controls (*\*\*\*\*p<0.0001*; PD: mean % area: 1.58 %, 95% CI 1.40 to 1.75; HCs: mean % area: 0.18 %, 95% CI 0.09 to 0.26) (Figure 1P).

**Figure 1.**
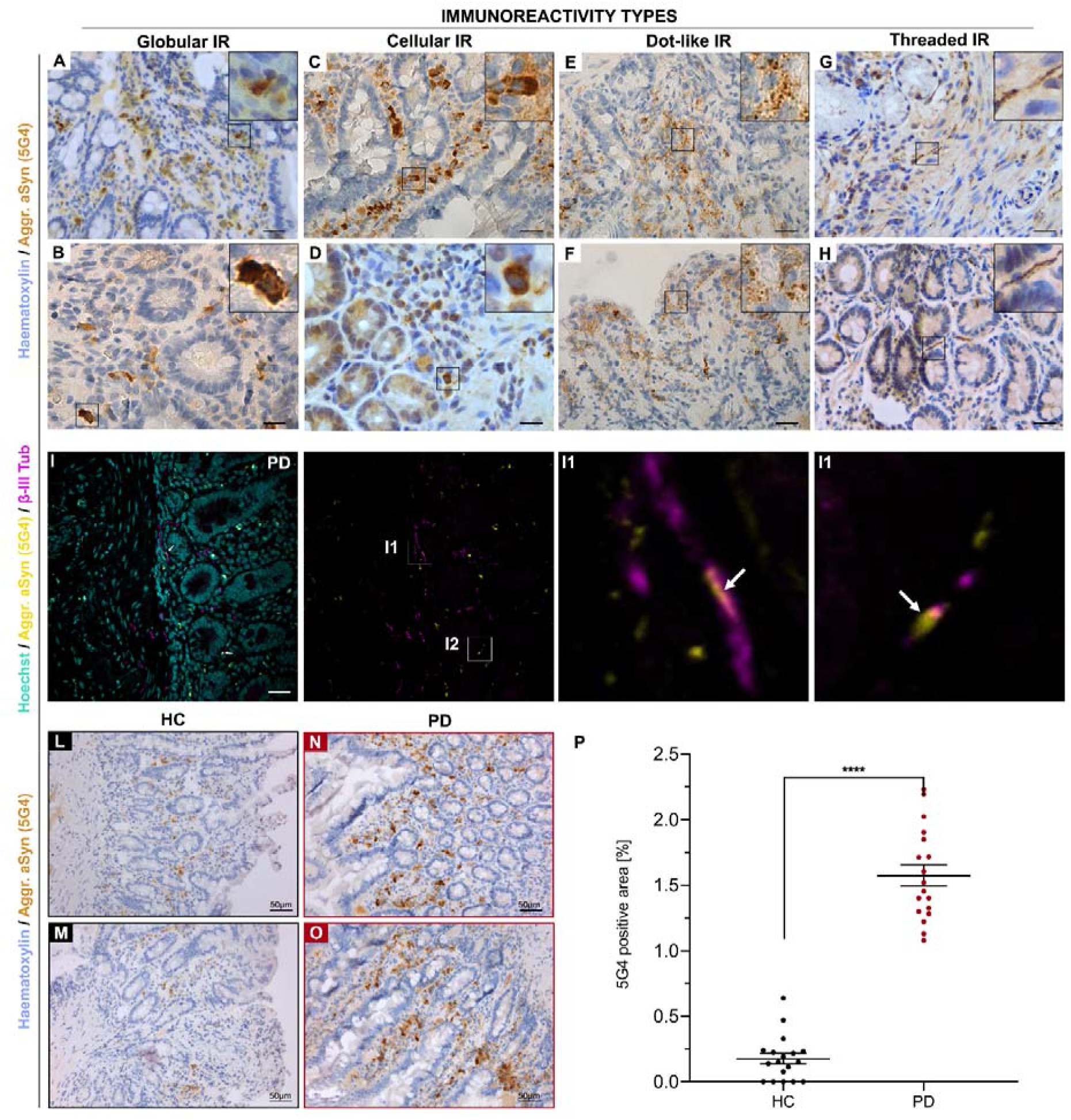
αSyn-5G4 immunohistochemistry in duodenal biopsies. A-I) 5G4 Immunoreactivity types encountered in the duodenal mucosa. A-B) Globular immunoreactivities and C-D) cellular reactivities, found in both Parkinson’s Disease patients (PD) and Healthy Controls (HC), likely represent antibody cross-reactivity with resident mast cells. E-F) Dot-like reactivities can be either ascribed to lipopigment deposits in the GI mucosa, or represent actual 5G4 aggregated synuclein deposits in nerve fibers fragmented within the sectioning plane. G-H) Threaded (thread-like) reactivities encountered predominantly in PD patients indicate alpha-synuclein aggregates in nerve fibers of the GI tract, as demonstrated by double-label immunofluorescent staining for 5G4 aggregated synuclein (yellow) and beta-III-tubulin (magenta), a pan-neuronal and neuritic marker (I1-I2). Overall, αSyn-5G4 immunoreactivity was lower in HC (L-M) compared to PD (N-O). Morphometrical quantification followed by Mann-Whitney U-test reveals statistically significant differences between PD and HC (*\*\*\*\*p<0.0001*) (P).

Next, we defined the criteria for diagnosing PD-related αSyn pathology with two parameters: 1) an average morphometric area of aggregated αSyn burden in at least three counting fields equal or higher than 0.26 % (upper 95% CI of mean in HCs) of the total counting field (A% > 0.26) and/or 2) at least a single unequivocal thread-like profile detected by αSyn-5G4.

Application of parameter Nr. 1 alone revealed a sensitivity of 100.00% (95%CI=81.47-100.00%), specificity of 83.33% (95%CI=58.58-96.42%), Positive Predictive Value of 85.71% (95%CI=68.11-94.40%) and Negative Predictive Value of 100% for discerning manifest PD from HC. The calculated accuracy was thus equal to 91.67% (95%CI=77.53-98.25%).

Application of parameter Nr. 2 alone revealed a sensitivity of 100.00% (95%CI=81.47-100.00%), specificity of 94.44% (95%CI=72.71-99.86%), Positive Predictive Value of 94.74% (95%CI=72.82-99.18%) and Negative Predictive Value of 100.00% for discerning manifest PD from HC. The calculated accuracy was thus equal to 97.22% (95%CI=85.47-99.93%) and was higher than parameter Nr. 1.

Combination parameters Nr. 1 and 2 revealed a sensitivity of 100.00% (95%CI=81.47-100.00%), specificity of 100.00% (95%CI=81.47-100.00%), Positive Predictive Value of 100.00% and Negative Predictive Value of 100.00% for discerning manifest PD from HC. The calculated accuracy was thus equal to 100.00% (95%CI=85.47-99.93%) and was higher than the single parameters.

### GFAP analysis and enteric gliosis

GFAP immunoreactive elements presented as discrete, round immunoreactive cells localized predominantly within the duodenal mucosa and submucosa (Figure 2A, 2H High-Magnification Inserts).

**Figure 2.**
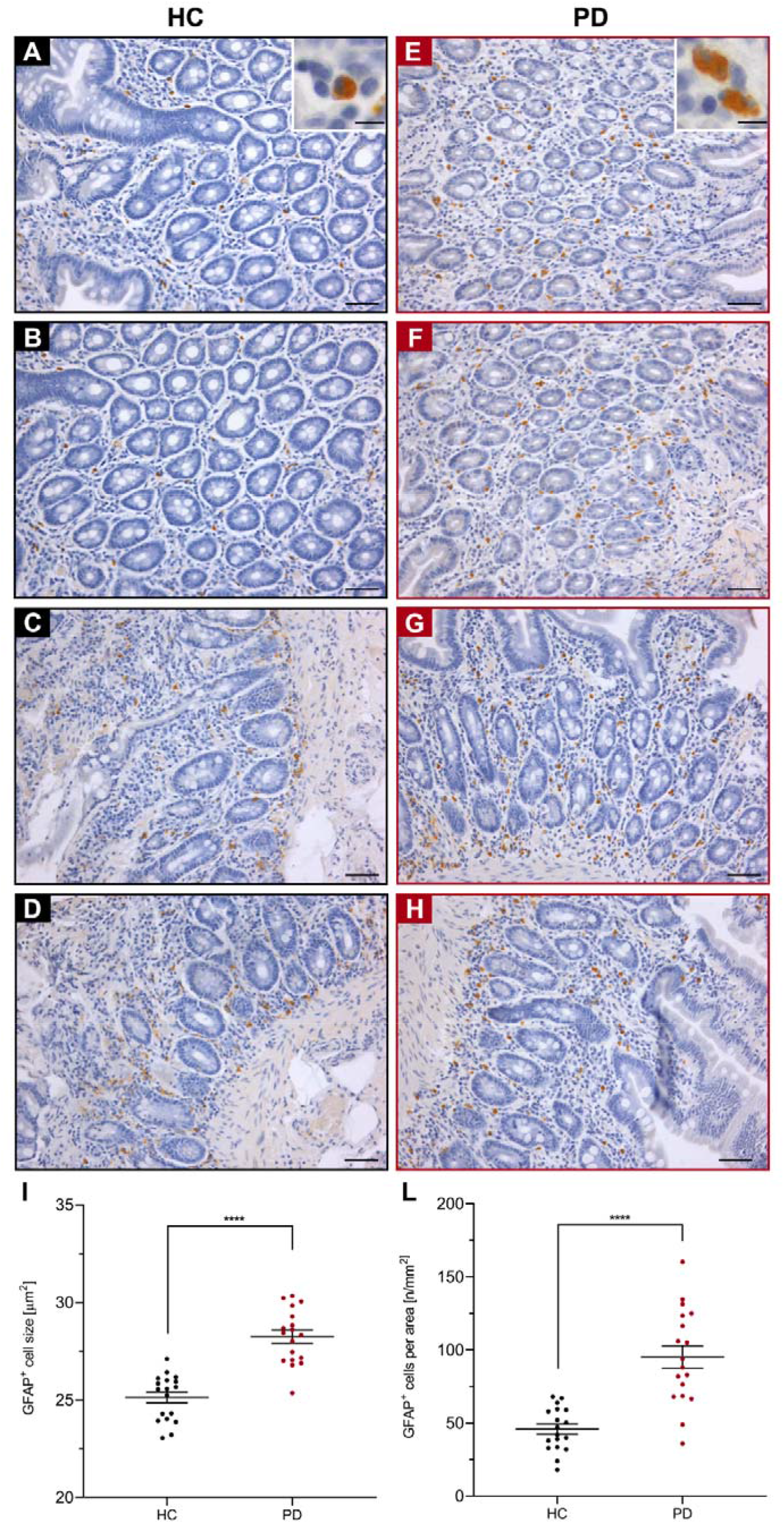
GFAP immunoperoxidase-stained duodenal biopsies. A-D) Healthy Controls (HC). E-H) Parkinson’s Disease (PD) patients. Scale Bar = 50µm; high magnification inserts Scale Bar = 15µm. I-L) Mann-Whitney U-test reveals statistically significant differences between PD and HC in terms of both enteric glial cell size (I) and density (L) (*\*\*\*p<0.001; ****p<0.0001*).

Morphometrical analyses revealed both increased EGC density (PD mean: 95.2±32.6; CTRL mean: 45.8±14.9; Figure 2L) and increased cell size (PD mean: 28.2±1.4 µm^2^; CTRL mean: 25.1±1.2 µm^2^; Figure 2I) in PD patients compared to HCs (*\*\*\*\*p<0.0001* for both cell density and size), suggesting for local reactive gliosis.

Spearman’s rank correlation analysis between αSyn-5G4 immunoreactive area, EGC density and EGC cell size in the duodenum revealed a strong correlation (R_s_=0.751, *p<0.0001*□and R_s_=0.741, *\*\*\*\*p<0.0001*, n=36, respectively).

### Correlation with clinical data

Pearson’s correlation analysis was performed between αSyn-5G4 immunoreactive areas and collected clinical data. No correlation was found with sex, disease duration, motor severity, global cognitive scales’ score, quality of life, and MDS-UPDRS scale score. Interestingly, we found a trend of increased aggregated αSyn distribution in patients who complained of constipation (*p*=0.058) (WCS), however validation of this finding requires a larger cohort as a prospective assessment with validated instruments. Similarly, no significant correlation was observed with the other variables examined, including other gastrointestinal disturbances.

## DISCUSSION

In the present study, we documented marked immunoreactivity for aggregated αSyn and morphological changes in EGC suggestive of reactive gliosis in the duodenum of advanced PD patients. These data expand our knowledge of the involvement of the enteric nervous system in PD and suggest the duodenum as possible target for early disease detection.

Our results also indicate that the αSyn-5G4 conformation specific αSyn antibody is a reliable marker for discerning duodenal biopsies of PD patients and HCs. While low or barely detectable immunoreactivity for αSyn-5G4 was also found in controls, marked differences with PD patients were revealed by morphometrical quantification. Furthermore, morphological evaluation revealed thread-like immunoreactivities as PD specific-findings; immunofluorescent staining confirmed colocalization between αSyn aggregates and neuronal markers, indicating synuclein aggregates in nerve fibers of the duodenal mucosa and submucosa. αSyn-5G4 antibody appears to be specific for αSyn aggregates, as monomeric synuclein is not detectable in vitro^24^. Furthermore, phospho-αSyn antibodies are not reliable to discern between PD patients and controls in gastric and colonic mucosa specimens, thus significantly limiting their usefulness^35 36^. Unlike previous studies, we have examined an anatomically defined region of the small intestine, the duodenum, greatly restricting the sampling site variability present in colonic mucosa studies. Moreover, the left half of the transverse colon and the descending colon receive parasympathetic innervation from the sacral nerves deriving from the spinal cord, and not from the vagus nerve, and thus are inherently biased and likely less relevant towards the brain-to-gut hypothesis of αSyn transmission. The localization and density of αSyn aggregates in the average aging population have not been documented to date, with low quantities of aggregated αSyn possibly representing a normal finding in aging subjects, as seen in this small cohort.

GFAP immunoreactive cell density and size were also higher in PD patients compared to HCs, suggesting enteric gliosis. However, immunofluorescent staining for αSyn-5G4 and GFAP antibodies did not reveal colocalization between markers (Supplementary Figure 1). Conversely, αSyn-5G4 colocalization with neuronal marker β-III-tubulin was found, indicating a preferential site for αSyn aggregates. Challis et al.^37^ evidenced increased myenteric EGCs volume and count in a mouse model of αSyn preformed-fibrils (PFF) duodenal inoculation, similar to our in-vivo findings. According to the authors, αSyn-PFF inoculation induces reactive gliosis in response to fibrils seeding, thus linking αSyn pathology to EGCs reaction. Our study suggests a localized response of EGCs in in-vivo PD patients in line with the current animal-model studies, which is also supported by the strong correlation between EGC values and aggregated αSyn; however, the mechanisms underlying this require further investigation in humans, with regard to inflammatory and immunity processes involved^38^.

There was no correlation between patients’ clinical characteristics, including cognitive scales and αSyn-5G4 distribution in the duodenum, suggesting that peripheral pathology may not necessarily reflect phenotypic variability. To investigate this hypothesis further, prospective studies could include REM Behavior Sleep Disorder (RBD), extensive cognitive assessment allowing cognitive status characterization as well as early PD subjects. Inclusions of patients at various disease stages may provide further insight into the temporal dynamics of aggregated αSyn in the GI tract and its association with brain pathology.

Limitations of this study include the small sample size and the lack of patients with non-severe disease stages. On the other hand, strength of this study includes the prospective and systematic assessment of the patients and the extensive recording of clinical data which made the correlations more reliable.

In conclusion, our data suggest that duodenal biopsy may represent a safe, feasible and valuable tool for characterizing PD pathology in the GI tract and discerning patients from controls. For diagnostic purposes we propose that the biopsy should be taken from a topographically unrelated site to PEG-J placement of the duodenal wall. For diagnostic evaluation a combination of a morphometric method (standardized for each laboratory) and detection of a single thread-like αSyn-5G4 immunoreactivity is recommended. Future studies will be required to confirm these findings in a prodromal or early PD phase and evaluate subjects with other synucleinopathies in particular multiple system atrophy.

## Data Availability

All data produced in the present study are available upon reasonable request to the authors

## Acknowledgements

The project was supported by ‘Segala award’ from SIN and Limpe to MiC. We thank all the donors and their families.

## Author Contributions

AA, GGK, LB, AE, MS designed the study. AA, FG, MiC, MaC recruited the patients. AA, MiC, MaC, FG performed the neurological evaluation of the participants. RB performed the neuropsychological assessment of the participants. FPR performed the endoscopic exam and sampling of the biopsies. AE, MS, GT performed the immunohistochemical staining of the samples. AE, MS, RDC, AP performed the morphometrical and morphological evaluation of immunoreactivities. AE, MS performed the statistical analyses. AE, MS, GGK, AA drafted the manuscript and figures. All authors read and approved the final version of the manuscript. All other authors declare no conflict of interest.

