## Supplementary material for "Duodenal alpha-Synuclein pathology and enteric gliosis in advanced Parkinson’s Disease patients": Supp. Table 1 and Supp. Fig. 1

**Supplementary Table 1**

| | Healthy controls N=18 | | PD patients N=18 | |
|---|---|---|---|---|
| | Median | 2.5 - 97.5 P | Median | 2.5 - 97.5 P |
| **AGE AT BIOPSY** | 68.5 | 54.0 - 86.0 | 66 | 48.0 - 79.0 |
| **SEX (M%)** | 50% | | 66.70% | |
| **AGE AT DIAGNOSIS** | | | 55.5 | 34.0 - 67.0 |
| **AGE AT LCIG INITIATION** | | | 65 | 45.0 - 76.0 |
| **YEARS OF DISEASE** | | | 12.5 | 3.0 - 21.0 |
| **LEDD AT LCIG INITIATION** | | | 1246.5 | 750.0 - 2588.0 |
| **MDS-UPDRS part I** | | | 10 | 6.0 - 24.0 |
| **MDS-UPDRS part II** | | | 17 | 2.0 - 37.0 |
| **MDS-UPDRS part III** | | | 32.5 | 10.0 - 58.0 |
| **MDS-UPDRS part IV** | | | 8 | 3.0 - 13.0 |
| **H&Y>2** | | | 43.80% | |
| **PDQ-8** | | | 12 | 0.0 - 21.0 |
| **ADL** | | | 5 | 2.0 - 6.0 |
| **IADL** | | | 5 | 2.0 - 8.0 |
| **PD-CFRS** | | | 1 | 0.0 - 9.0 |
| **MMSE (corrected score)** | | | 26.2 | 22.0 - 30.0 |
| **MoCA (corrected score)** | | | 24.52 | 17.1 - 30.0 |

***Supp. Table 1.*** *Demographic and clinical characteristics of Parkinson's Disease and healthy controls patients as median ± standard deviations and 97,5% confidence interval (CI) are shown. PD: Parkinson's Disease. LCIG levodopa/carbidopa intestinal gel, MMSE Mini-Mental State Examination, MOCA Montreal Cognitive Assessment, PDQ8 Parkinson's Disease Questionnaire, MDS-UPDRS Unified Parkinson's Disease Rating Scale, H&Y Hoehn-Yahr Staging Scale, LEDD Levodopa Equivalent Dose Calculator, IADL impairment of instrumental activities of daily living, ADL impairment of activities of daily living.*

**Supplementary Figure 1. Aggregated αSyn (5G4 antibody) do not colocalize with enteric glia in duodenum biopsies.**

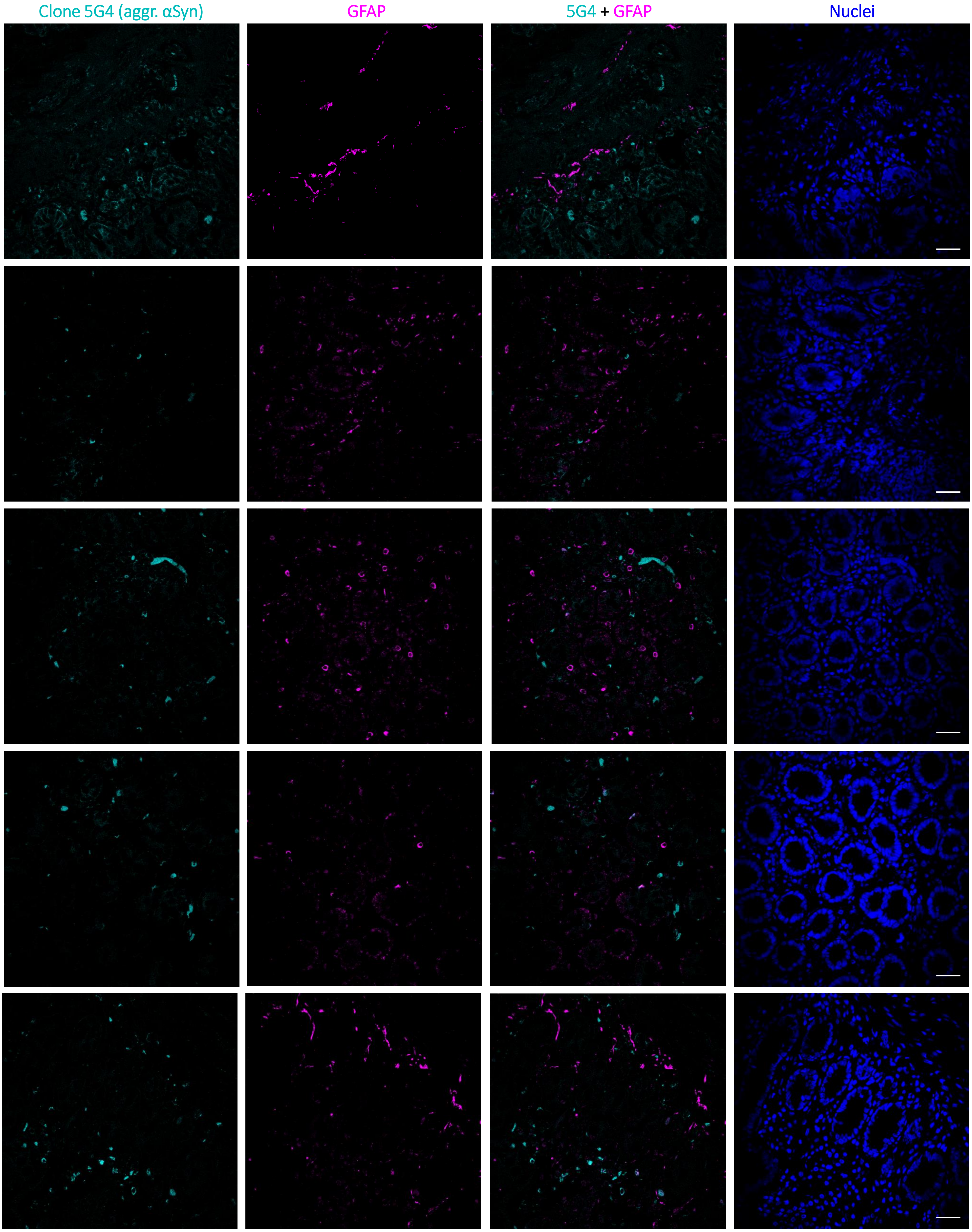


***Supp. Fig 1. Aggregated αSyn (5G4 antibody) do not colocalize with enteric glia in duodenum biopsies.*** *Representative immunofluorescence images of duodenal biopsies from PD patients. Both GFAP (magenta), the cellular glial marker, and aggregated α-Syn (cyan) are visible but are not colocalizing as white spot signals. Scale bar: 60 µm.*
